# Glucagon-Like Peptide 1 Receptor Agonist Use in Hospital: A Multicentre Observational Study

**DOI:** 10.1101/2024.08.01.24311352

**Authors:** Prachi Ray, Jason A. Moggridge, Alanna Weisman, Mina Tadrous, Daniel J. Drucker, Bruce A. Perkins, Michael Fralick

## Abstract

**Introduction:** Glucagon-like peptide 1 receptor agonists (GLP-1RA) are effective medications for type 2 diabetes mellitus (T2DM) and obesity, yet their uptake among patients most likely to benefit has been slow.

**Methods:** We conducted a cross-sectional analysis of medication exposure in adults hospitalized at 16 hospitals in Ontario, Canada between 2015 and 2022. We estimated the proportion with T2DM, obesity, and cardiovascular disease. We identified the frequency of GLP-1RA use, and conducted multivariable logistic regression to identify factors associated with their use.

**Results:** Across 1,278,863 hospitalizations, 396,084 (31%) patients had T2DM and approximately 327,844 (26%) had obesity. GLP-1RA use (n=1,274) was low among those with T2DM (0.3%) and those with obesity (0.7%), despite high prevalence of cardiovascular disease (36%). In contrast, use of diabetes medications lacking cardiovascular benefits was high, with 60% (n=236,612) receiving insulin and 14% (n=54,885) receiving sulfonylureas. Apart from T2DM (OR=29.6, 95% CI 23.5, 37.2), characteristics associated with greater odds of receiving GLP-1RA were age 50-70 years (OR=1.71, 95% CI 1.38, 2.11) compared to age < 50 years, hemoglobin A1C > 9% (OR=1.83, 95% CI 1.36, 2.47) compared to < 6.5%, and highest income quintile (OR=1.73, 95% CI 1.45, 2.07) compared to lowest income quintile.

**Conclusion:** Knowledge translation interventions are needed to address the low adoption of GLP-1RA among hospitalized patients with T2DM and obesity, who are the most likely to benefit.

## Introduction

Glucagon-like peptide 1 receptor agonists (GLP-1RA) are effective medications for adults with obesity or type 2 diabetes mellitus (T2DM).^1–4^ Multiple randomized controlled trials (RCTs) have shown that GLP-1RA reduce the relative risk of cardiovascular events (i.e., myocardial infarction, stroke, and cardiovascular death) by approximately 15% in adults with T2DM, and by 20% in adults with cardiovascular disease and obesity but without T2DM.^3,5^ GLP-1RA lead to reductions in body weight of approximately 5-10% in adults with T2DM and approximately 15% in adults without T2DM who are overweight or living with obesity.^2,6^ There is a 24% relative risk reduction of a primary outcome encompassing end-stage renal disease and cardiovascular disease in adults with T2DM treated with GLP-1RA.^7^ GLP-1RA lead to a two-fold reduction in heart-failure symptoms in people with heart failure with preserved ejection fraction, for those with either T2DM and obesity, or obesity alone.^8^ Clinical guidelines now recommend GLP-1RA as one of the two recommended second-line treatments for adults with T2DM.^9^ For adults with obesity and obesity-related co-morbidities, GLP-1RA are the first-line pharmacotherapy treatment.^10^

Despite guidelines consistently recommending their use, uptake of GLP-1RA among patients most likely to benefit from them remains low. This is concerning given the high prevalence of diabetes and obesity in North America.^11,12^ Systematic reviews have identified common barriers to prescribing newer medications for chronic disease, including both patient and provider-level barriers.^13,14^ Patients may be hesitant because of potential side effects, costs, and route of administration, all of which are directly relevant to GLP-1RA. In Canada, the cost of a one-month supply for adults without medication coverage is approximately $200 USD; in the United States, it can cost up to $1000 USD per month.^15^ Ideally, preferential use should be for patients who are most likely to benefit, rather than those most likely to afford the medications. The available clinical trials demonstrate that the patients most likely to benefit from GLP-1RA are older adults with cardiovascular disease and either T2DM or obesity.^1–3,5,7,8^ However, the high out-of-pocket cost of GLP-1RA for adults without insurance has led to their preferential use among individuals with higher socioeconomic status.^16^

Hospitalized adults are one of the highest risk groups for subsequent cardiovascular events, because inpatients have a median age of 73 years, have a median of six underlying chronic medical conditions, and the reason for the hospitalization can directly (e.g., hospitalization for myocardial infarction) or indirectly (e.g., influenza, COVID-19) increase a person’s risk of subsequent cardiovascular events.^17–19^ Starting new medications for chronic diseases in-hospital, including at the time of discharge, leads to better medication adherence compared to deferring the decision to the outpatient setting.^20,21^ Medication adherence is especially important for T2DM and obesity because both are chronic diseases requiring long-term treatment. Therefore, the inpatient setting presents a valuable opportunity to start patients on medications such as GLP-1RA. Our primary objective was to determine the frequency of, and factors associated with, GLP-1RA use among hospitalized adults, who represent a high-risk population poised to benefit from GLP-1RA.

## Methods

### Data Source

We conducted a cross-sectional analysis to determine the frequency of GLP-1RA use across 16 hospitals in Ontario, Canada, using retrospective data from the General Medicine Inpatient Initiative (GEMINI) cohort. GEMINI represents the largest repository of inpatient data in Canada, and data within GEMINI have an accuracy of > 98%.^22^ Within GEMINI, patients’ medical records are linked to administrative datasets including Canadian Institute for Health Information (CIHI) and Discharge Abstract Database (DAD). GEMINI includes data on discharge diagnosis, patient demographics (e.g., age, sex, place of residence), laboratory results (e.g., HbA1C), imaging reports, vital status at discharge (e.g., death), medications, and other administrative data (e.g., length of stay). GEMINI also includes a proxy for socioeconomic status (SES), determined through postal code, which can be used to assess neighbourhood-level SES through linkage with Canadian census data (i.e., household income levels). Quintile 1 is the lowest income range while quintile 5 is the highest income range.^23^ Data not available within GEMINI include physician-level data (e.g., specialty, gender), nursing notes, or daily charting notes.

### Study Population

We identified hospitalizations of patients over 18 years of age who were admitted to general internal medicine between 2015 to 2022 (most recent available data). Of these hospitalizations, we identified those in which a GLP-1RA was administered. We identified hospitalizations with T2DM using International Classification of Diseases, Tenth Revision (ICD-10) diagnosis codes for T2DM in any of the following fields: current or past admission; main responsible diagnosis; pre-admit, secondary, admitting, or transfer diagnosis. The ICD-10 codes for T2DM were recently validated within GEMINI (e.g., PPV and sensitivity > 95%).^24^ We identified hospitalizations with obesity using previously validated ICD-10 codes which are specific but not sensitive (specificity > 99%, sensitivity = 9%).^25^

### Study Objectives

The primary objective was to identify the frequency of GLP-1RA use in hospital and the factors associated with their use. Given the cardioprotective benefits of GLP-1RA, we also reported the proportion of hospitalizations with T2DM and the following comorbidities using ICD-10 codes which have been previously validated in administrative databases: coronary artery disease (CAD), stroke or transient ischemic attack (TIA), peripheral vascular disease (PVD), heart failure, and renal disease.^9,26–29^ We were unable to estimate the number of hospitalizations with obesity and cardiovascular disease because of the low sensitivity of ICD-10 codes for obesity. Our study received Research Ethics Board approval from the participating hospitals.

### Sensitivity Analysis

We conducted a sensitivity analysis of GLP-1RA use in the last complete calendar year of our study (i.e., January 1, 2021 - January 1, 2022). This was done to assess the robustness of our study findings, because there may be time-varying factors (e.g., availability of GLP-1RA in hospital) contributing to GLP-1RA use between the 2015-2022 study period which were not accounted for in our primary analysis. We identified a cohort of all patients with T2DM who were admitted to the hospital in 2021 and reported the percentage use of diabetes medications among this cohort. Among those who did not receive GLP-1RA, we identified the number of times they were hospitalized in the preceding six years and reported the mean count of hospitalizations.

### Statistical Analysis

Descriptive statistics were used to compare the patient-level characteristics of hospitalizations in which a GLP-1RA was administered in hospital compared with hospitalizations who did not, and we reported the standardized mean difference (SMD) between the two groups. The accepted cut-off of an SMD is 0.10; if the SMD is above 0.10, the two groups are considered imbalanced.^30^ We identified hospitalizations with T2DM and obesity using ICD-10 codes. Given the low sensitivity of ICD-10 codes for obesity (i.e., 9%), we multiplied the number of hospitalizations with an ICD-10 code for obesity by 100%/9% to estimate the total number of patients with obesity within GEMINI.

To identify factors associated with GLP-1RA use, we built a multivariable logistic regression model. The model included the following variables, which were selected based on content expertise and prior literature: demographics (i.e., age, sex, postal code [as a proxy for SES]), comorbidities (i.e., T2DM, obesity, cardiovascular disease, dementia, hypertension, dyslipidemia, renal disease), and results from inpatient laboratory tests (i.e., HbA1C, creatinine).^7^ We also included the number of years since 2015, and each individual hospital (anonymized through identification numbers) as variables in this model.^13^ Odds ratio estimates and 95% Wald confidence intervals were reported. All statistical analyses were performed using the R Statistical Software v4.0.5 (R Foundation for Statistical Computing, Vienna, Austria).

## Results

### Study Population

We identified a total of 1,278,863 hospitalizations between 2015 and 2022. The median age was 70 years (IQR 56.0-82.0), 47% were female, and the three most common discharge diagnoses were heart failure, pneumonia, and chronic obstructive pulmonary disease/asthma (Table 1). Overall, 40% of hospitalizations had hypertension, 22% had CAD, 8% had dyslipidemia, 10% had heart failure, and 13% had renal disease. Median HbA1c was 6.1% (IQR 5.5-7.4) and median creatinine was 87 μmol/L (IQR 68.0-123.0) at baseline for these hospitalizations.

**Table 1.**
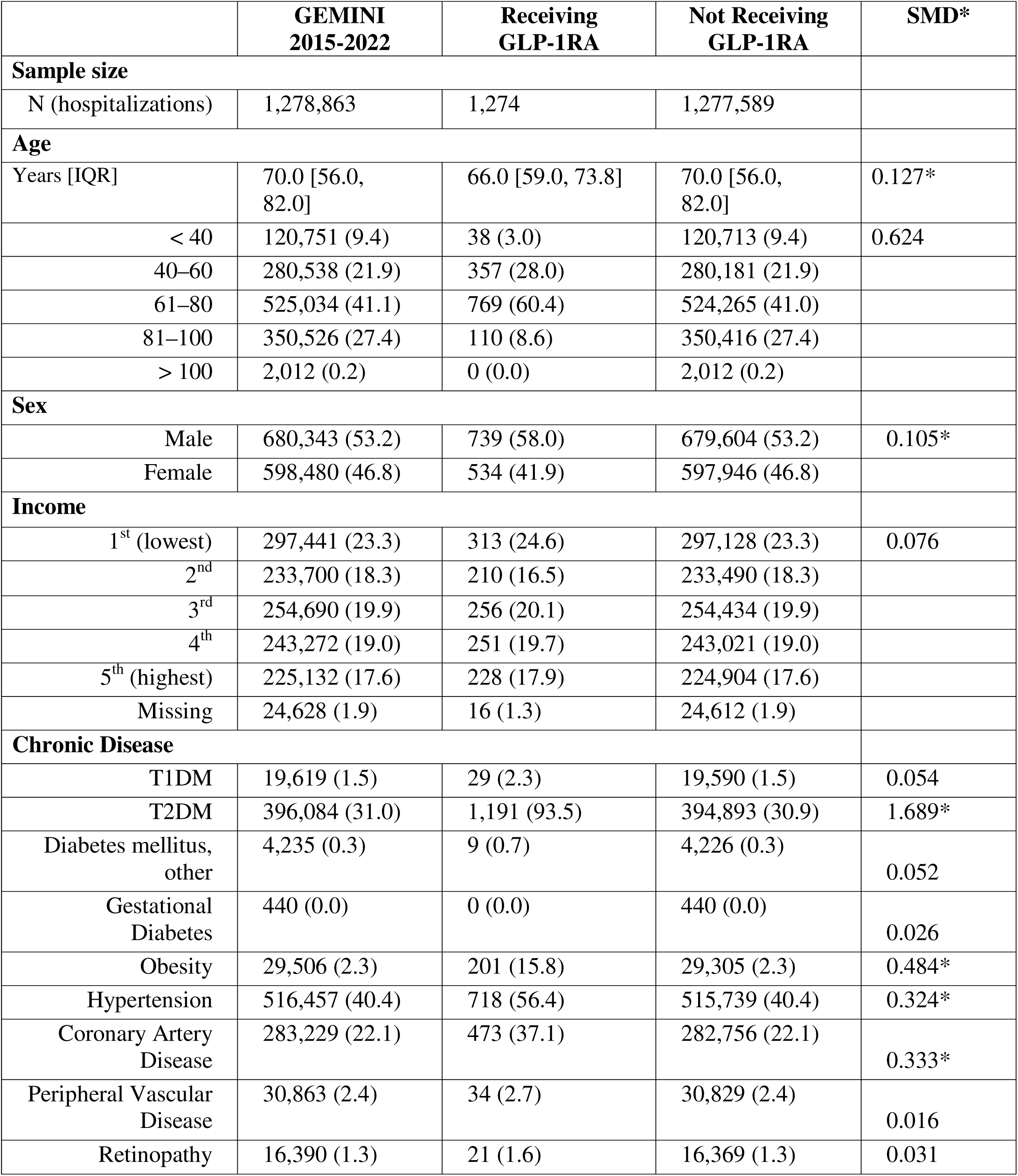
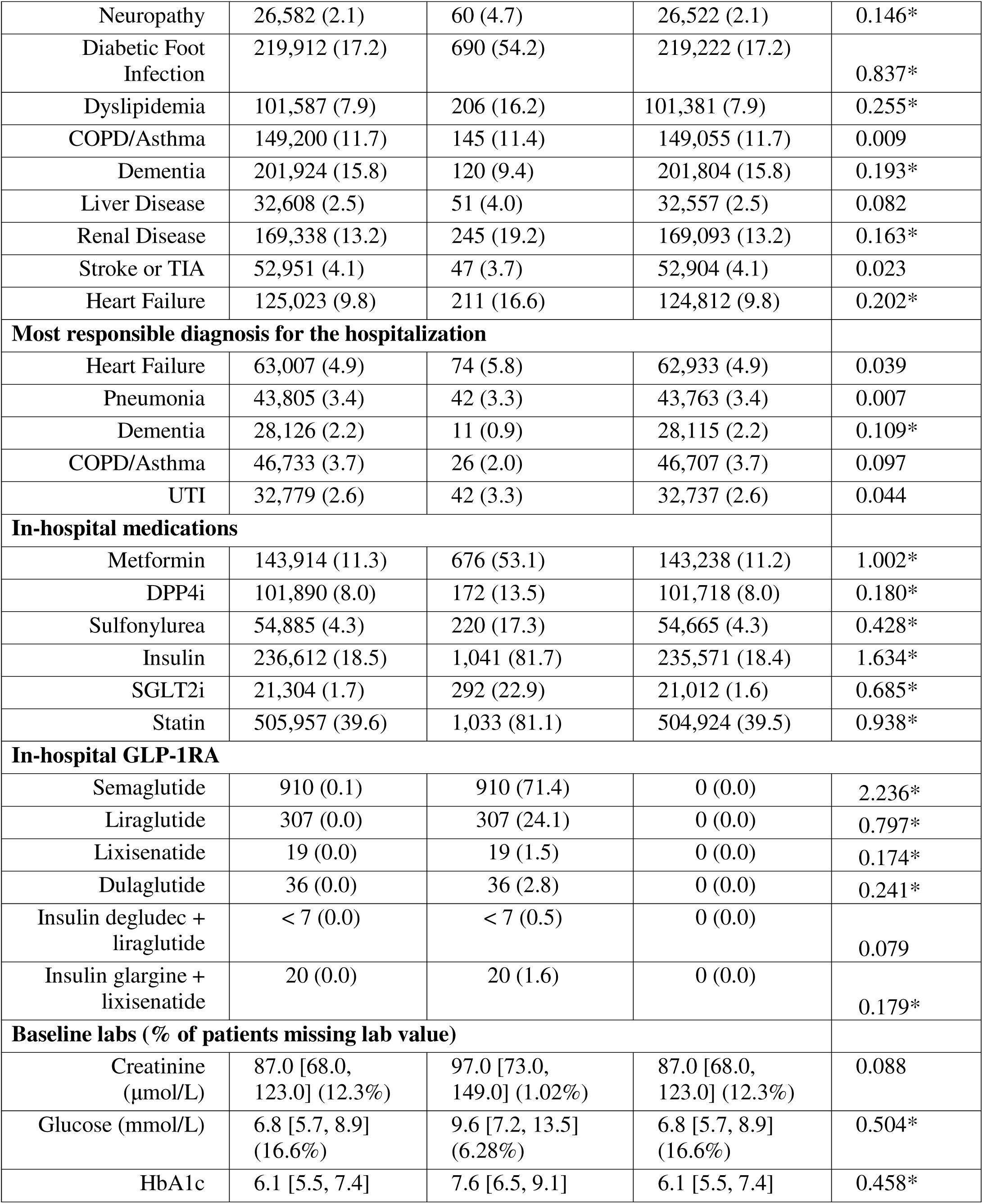
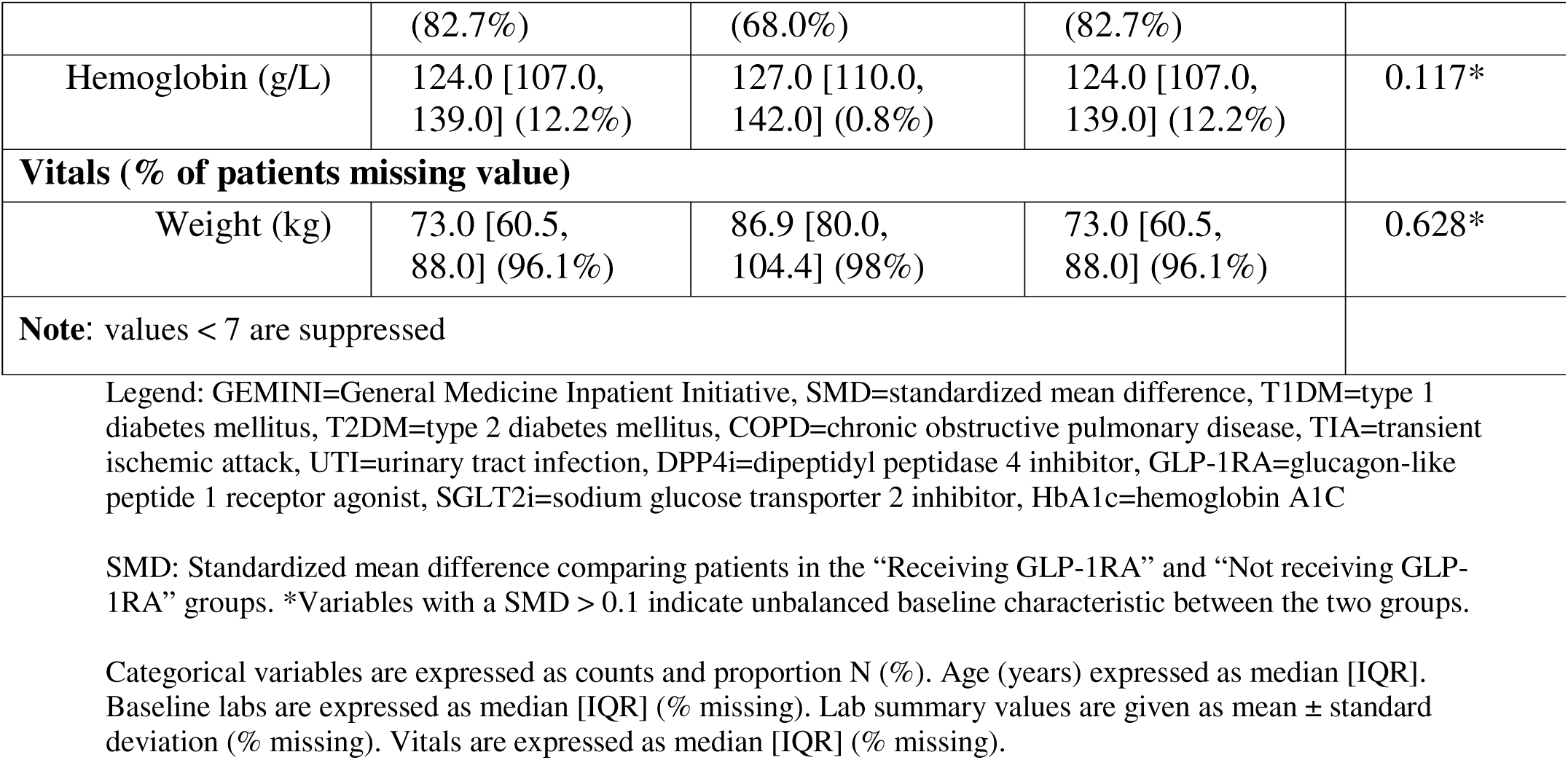
Demographic and clinical characteristics associated with hospitalizations.

We identified 1274 (0.1%) hospitalizations in which a GLP-1RA was administered. Patients who received a GLP-1RA were younger (66 years vs 70 years), more likely to be male (58%), and had a higher prevalence of T2DM (94% vs 31%) or obesity (16% vs 2%) compared to those who did not receive a GLP-1RA (Table 1). Patients who received a GLP-1RA had a higher prevalence of hypertension (56% vs 40%), CAD (37% vs 22%), and dyslipidemia (16% vs 8%) compared to patients who did not receive a GLP-1RA.

### Type 2 Diabetes Mellitus

One-third of all hospitalizations involved patients with T2DM (n=396,084). Of these, 1,191 hospitalizations (0.3%) included administration of a GLP-1RA. In contrast, metformin was administered in 36% (n=143,914) of the hospitalizations, insulin in 60% (n=236,612), sulfonylurea in 14% (n=54,885), dipeptidyl peptidase 4 inhibitor (DPP-4i) in 26% (n=101,890), and sodium-glucose co-transporter-2 inhibitor (SGLT2i) in 5% (n=21,304). Among hospitalizations with T2DM, 31% (n=123,708) had CAD, 6% (n=24,003) had stroke or TIA, 3% (n=10,063) had PVD, 17% (n=66,366) had heart failure, and 22% (n=88,494) had renal disease (Table 2). Approximately 36% (n=143,547) of patients with T2DM had cardiovascular disease (i.e., CAD, stroke or TIA, or PVD).

**Table 2.** Number of patients with Type 2 Diabetes Mellitus who would benefit from GLP-1RA.

| **History of Condition** | **N encounters (%) in Hospitalizations with T2DM** |
| --- | --- |
| Coronary Artery Disease (CAD) | 123,708 (31.2%) |
| Stroke or Transient Ischemic Attack (TIA) | 24,003 (6.1%) |
| Peripheral Vascular Disease (PVD) | 10,063 (2.5%) |
| Heart Failure | 66,366 (16.8%) |
| Renal Disease | 88,494 (22.3%) |
| CAD, Stroke, or TIA | 138,689 (35.0%) |
| CAD, Stroke, TIA, or PVD | 143,547 (36.2%) |
| CAD, Stroke, TIA, PVD, or heart failure | 170,419 (43.0%) |
| CAD, Stroke, TIA, PVD, heart failure, or renal disease | 201,586 (50.9%) |

Legend: This represents the number of hospitalizations associated with the above conditions. There were a total of 396,084 hospitalizations with Type 2 Diabetes Mellitus.

### Obesity

Overall, 2% (n=29,506) of all hospitalizations had an ICD-10 diagnosis code for obesity. Weight data were available for 4% (n=51,154) of patients. The median weight for patients on GLP-1RA was 87 kg (IQR 80-104) and the median weight for patients not on GLP-1RA was 73 kg (IQR 61-88). Because the ICD-10 codes for obesity have a sensitivity of 9%, we estimated that the true prevalence of obesity was 327,844 (i.e., 29,506 x 100%/9%), corresponding to approximately 26% of patients. We did not report comorbidities among patients with obesity because the number of hospitalizations in our data with an ICD-10 code for obesity was low. Our point prevalence study of 35 patients on the general inpatient wards at Mount Sinai Hospital identified that 26% (n=9) of patients were obese (BMI ≥ 30).

### Multivariable Logistic Regression Model

In our multivariable logistic regression model, the factor most strongly associated with receiving a GLP-1RA was T2DM (Odds Ratio (OR)=29.6, 95% Confidence Interval (CI) 23.5, 37.2) (Table 3). The following factors were also associated with higher odds of receiving GLP-1RA: obesity (OR=2.4, 95% CI 2.04, 2.83), age 50-70 years (OR=1.71, 95% CI 1.38, 2.11) compared with age < 50 years, and dyslipidemia (OR=1.28, 95% CI 1.07, 1.54). Those with HbA1c between 6.5-9% (OR=1.67, 95% CI 1.28, 2.18) and HbA1c > 9% (OR=1.83, 95% CI 1.36, 2.47) were more likely to be prescribed GLP-1RA, compared with individuals with HbA1c < 6.5%. Creatinine levels between 100-200 μmol/L (OR=1.18, 95% CI 1.04, 1.35) were associated with increased odds of receiving GLP-1RA compared to creatinine levels < 100 μmol/L.

**Table 3.**
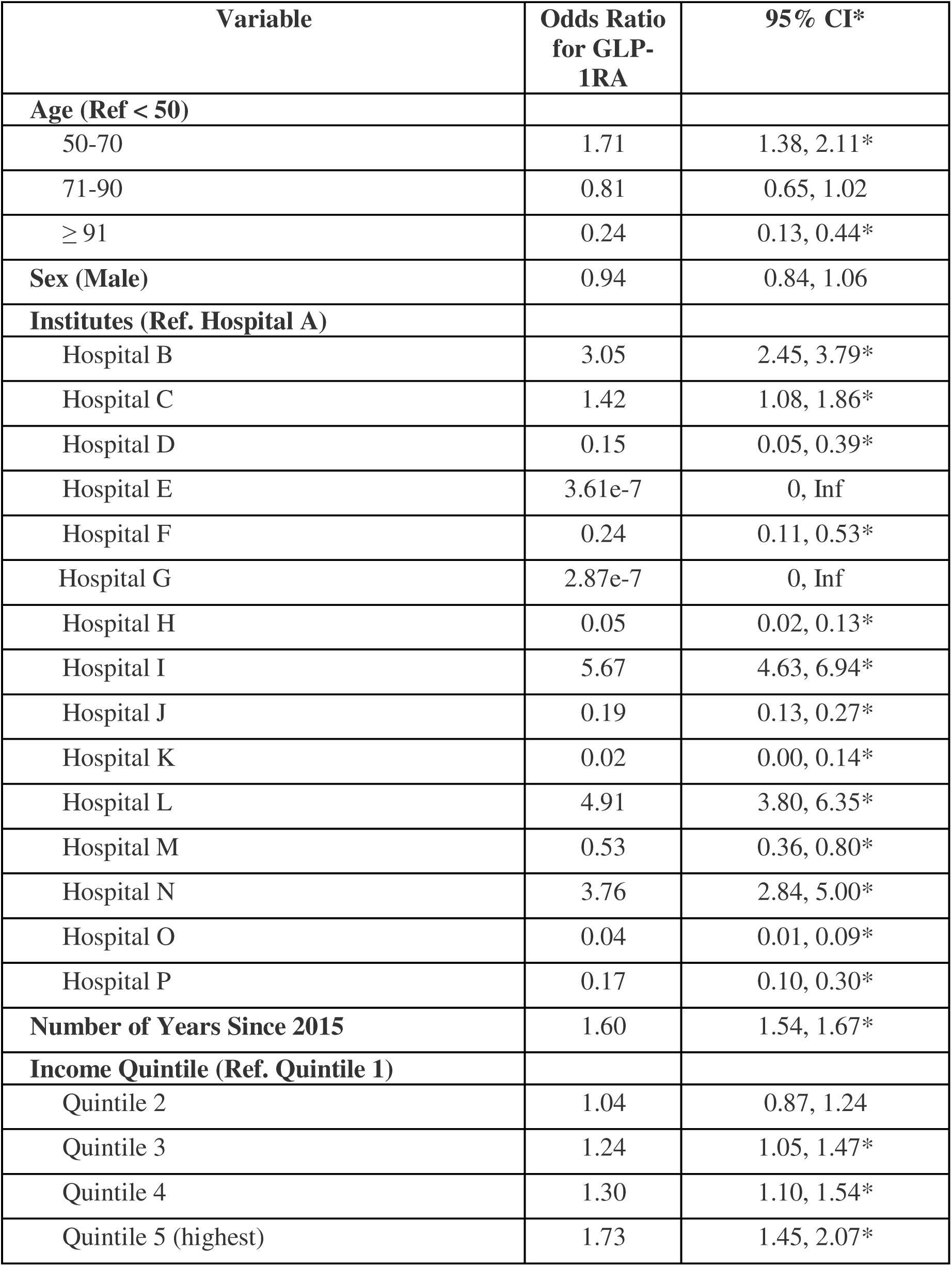
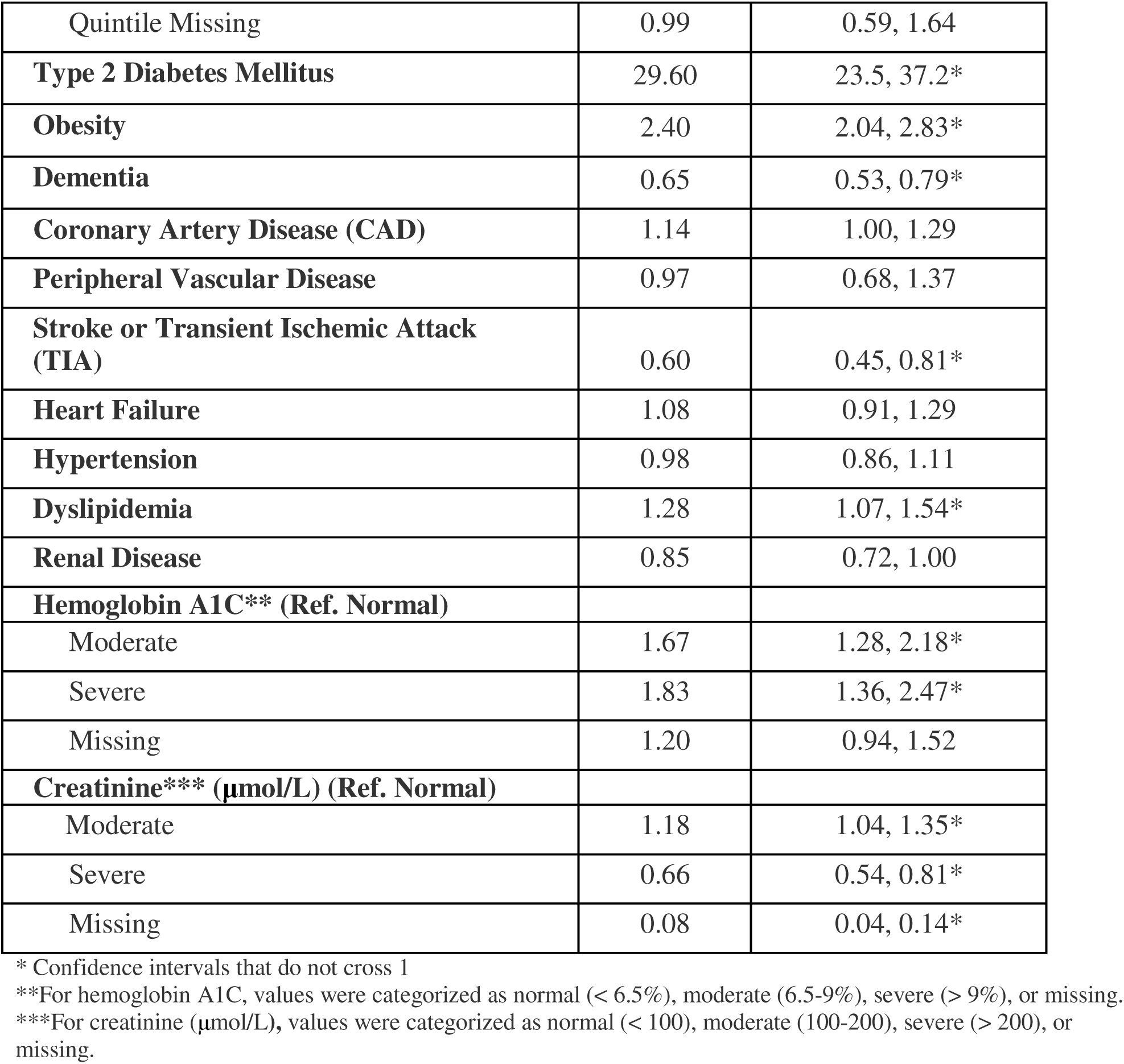
Multivariable logistic regression model identifying factors associated with GLP-1RA use in hospital.

| Variable | Odds Ratio<br>for GLP-1RA | 95% CI* |
| --- | --- | --- |
| **Age (Ref < 50)** |  |  |
| 50-70 | 1.71 | 1.38, 2.11* |
| 71-90 | 0.81 | 0.65, 1.02 |
| ≥ 91 | 0.24 | 0.13, 0.44* |
| **Sex (Male)** | 0.94 | 0.84, 1.06 |
| **Institutes (Ref. Hospital A)** |  |  |
| Hospital B | 3.05 | 2.45, 3.79* |
| Hospital C | 1.42 | 1.08, 1.86* |
| Hospital D | 0.15 | 0.05, 0.39* |
| Hospital E | 3.61e-7 | 0, Inf |
| Hospital F | 0.24 | 0.11, 0.53* |
| Hospital G | 2.87e-7 | 0, Inf |
| Hospital H | 0.05 | 0.02, 0.13* |
| Hospital I | 5.67 | 4.63, 6.94* |
| Hospital J | 0.19 | 0.13, 0.27* |
| Hospital K | 0.02 | 0.00, 0.14* |
| Hospital L | 4.91 | 3.80, 6.35* |
| Hospital M | 0.53 | 0.36, 0.80* |
| Hospital N | 3.76 | 2.84, 5.00* |
| Hospital O | 0.04 | 0.01, 0.09* |
| Hospital P | 0.17 | 0.10, 0.30* |
| **Number of Years Since 2015** | 1.60 | 1.54, 1.67* |
| **Income Quintile (Ref. Quintile 1)** |  |  |
| Quintile 2 | 1.04 | 0.87, 1.24 |
| Quintile 3 | 1.24 | 1.05, 1.47* |
| Quintile 4 | 1.30 | 1.10, 1.54* |
| Quintile 5 (highest) | 1.73 | 1.45, 2.07* |
| Quintile Missing | 0.99 | 0.59, 1.64 |
| **Type 2 Diabetes Mellitus** | 29.60 | 23.5, 37.2* |
| **Obesity** | 2.40 | 2.04, 2.83* |
| **Dementia** | 0.65 | 0.53, 0.79* |
| **Coronary Artery Disease (CAD)** | 1.14 | 1.00, 1.29 |
| **Peripheral Vascular Disease** | 0.97 | 0.68, 1.37 |
| **Stroke or Transient Ischemic Attack (TIA)** | 0.60 | 0.45, 0.81* |
| **Heart Failure** | 1.08 | 0.91, 1.29 |
| **Hypertension** | 0.98 | 0.86, 1.11 |
| **Dyslipidemia** | 1.28 | 1.07, 1.54* |
| **Renal Disease** | 0.85 | 0.72, 1.00 |
| **Hemoglobin A1C\*\* (Ref. Normal)** |  |  |
| Moderate | 1.67 | 1.28, 2.18* |
| Severe | 1.83 | 1.36, 2.47* |
| Missing | 1.20 | 0.94, 1.52 |
| **Creatinine\*\*\* (μmol/L) (Ref. Normal)** |  |  |
| Moderate | 1.18 | 1.04, 1.35* |
| Severe | 0.66 | 0.54, 0.81* |
| Missing | 0.08 | 0.04, 0.14* |

\* Confidence intervals that do not cross 1
\*\*For hemoglobin A1C, values were categorized as normal (< 6.5%), moderate (6.5-9%), severe (> 9%), or missing.
\*\*\*For creatinine (μmol/L), values were categorized as normal (< 100), moderate (100-200), severe (> 200), or missing.

GLP-1RA use in hospital was also increasingly likely each year since 2015 (OR=1.6, 95% CI 1.54, 1.67), and patients had a higher odds of receiving GLP-1RA if they were admitted to certain institutions: hospital “B” (OR=3.05, 95% CI 2.45, 3.79), hospital “I” (OR=5.67, 95% CI 4.63, 6.94), hospital “L” (OR=4.91, 95% CI 3.8, 6.35), and hospital “N” (OR=3.76, 95% CI 2.84, 5). Patients belonging to a higher income quintile had higher odds of receiving GLP-1RA compared to patients in the lowest income quintile: quintile 3 (OR=1.24, 95% CI 1.05, 1.47), quintile 4 (OR=1.3, 95% CI 1.1, 1.54), and quintile 5 (OR=1.73, 95% CI 1.45, 2.07). Variables associated with lower odds of receiving GLP-1RA included older age (i.e., ≥ 91 years of age) (OR=0.24, 95% CI 0.13, 0.44) compared to younger age (i.e., < 50 years), dementia (OR=0.65, 95% CI 0.53, 0.79), renal disease (OR=0.85, 95% CI 0.72, 1), stroke or TIA (OR=0.60, 95% CI 0.45, 0.81), and creatinine greater than 200 μmol/L (OR=0.66, 95% CI 0.54, 0.81) compared to creatinine levels below 100 μmol/L.

### Sensitivity Analysis

Between January 1, 2021 to January 1, 2022, there were 38,040 patients with T2DM (Table 4). Among these 38,040 patients, use of GLP-1RA was 1% (n=407). In comparison, 62% (n=23,529) of patients received insulin and 13% (n=4,834) received a sulfonylurea. Among the 99% (n=37,633) of patients with T2DM who did not receive GLP-1RA, they had a mean number of three hospitalizations since 2015. Of these 37,633 patients, 24,197 had at least two hospitalizations during the previous six years, and 15,802 had at least three admissions since 2015.

**Table 4.** Number of patients with Type 2 Diabetes Mellitus between January 1, 2021 to January 1, 2022.

|  | Hospitalizations<br>Jan 1, 2021 - Jan 1, 2022<br>with T2DM | Number of unique patients<br>Jan 1 2021 - Jan 1, 2022<br>with T2DM |
| --- | --- | --- |
| Sample size | 54,647 | 38,040 |
| **In-hospital medications** |  |  |
| Metformin | 20,485 (37.5%) | 16,463 (43.3%) |
| DPP4i | 16,627 (30.4%) | 12,487 (32.8%) |
| Sulfonylurea | 5,889 (10.8%) | 4,834 (12.7%) |
| Insulin | 31,181 (57.1%) | 23,529 (61.9%) |
| SGLT2i | 6,159 (11.3%) | 5,218 (13.7%) |
| Statin | 34,794 (63.7%) | 25,590 (67.3%) |
| GLP-1RA | 480 (0.9%) | 407 (1.1%) |

Legend: T2DM=type 2 diabetes mellitus, DPP4i=dipeptidyl peptidase 4 inhibitor, GLP-1RA=glucagon-like peptide 1 receptor agonist, SGLT2i=sodium glucose transporter 2 inhibitor. Any hospitalization without a valid patient ID was excluded (n = 603).

## Discussion

In our multicentre retrospective study of over 1.2 million hospitalizations, use of GLP-1RA was rare (n=1,274) despite T2DM (n=396,084) and obesity (n=327,844) being common. Fewer than 0.5% of patients with T2DM or obesity received a GLP-1RA. Data from our study demonstrate a distinct gap between clinical evidence and clinical practice. GLP-1RA are a preferred second-line medication for T2DM, but we found that use of sulfonylureas was 43-fold higher and use of insulin was 185-fold higher than GLP-1RA use. Sulfonylureas and insulin have no cardiovascular benefits, and cause weight gain and hypoglycemia.^31^ While SGLT2i have established cardiovascular benefits for adults with T2DM, use of SGLT2i was also low in our study. Starting GLP-1RA while a patient is acutely unwell may be challenging, but this does not preclude its use before discharge when the patient has recovered, especially if the reason for the hospitalization is due to worsening glycemic control.^32^ Starting medications in hospital or prescribing them at discharge instead of deferring the decision to the outpatient setting leads to improved medication adherence and better management of chronic conditions.^20,21^

The 99% of patients with T2DM who were not on GLP-1RA in 2021 had an average of three hospitalizations in the past six years, indicating that despite multiple interactions with the healthcare system, they did not receive a GLP-1RA. While T2DM was the strongest factor associated with GLP-1RA use, the specific hospital to which a patient was admitted was strongly associated with both higher and lower odds of a patient receiving GLP-1RA. This finding is concerning, as care should ideally be consistent across hospitals, and patient-level characteristics should be the strongest factors associated with GLP-1RA use. Our study showed that comorbidities such as heart failure were not associated with GLP-1RA use in hospital, and stroke was paradoxically associated with lower odds of receiving GLP-1RA.

Explanations for the low use of GLP-1RA are likely multifactorial and related to clinician-level, patient-level, and system-level factors. Prescribing inertia is a common reason for slow uptake of new medications for chronic disease.^33–35^ Cost is also a likely factor limiting the broad uptake of GLP-1RA. In our study, patients living in areas of higher SES were more likely to receive a GLP-1RA compared to those living in lower SES areas, a finding consistent with studies of GLP-1RA use in the United States.^16,36^ In 2019, semaglutide was included in Ontario’s public coverage for T2DM treatment.^37^ For patients between the ages of 25-64 years who are not covered by the Ontario Drug Benefit program and do not have private insurance plans, paying out of pocket for GLP-1RA is a clear potential barrier.^38^ System-level factors, such as delays in adding GLP-1RA to the hospital formulary, may also have contributed to slow GLP-1RA uptake; each hospital in Ontario has its own decision-making process when adding medications to the hospital formulary.^39^

## Limitations

The GEMINI database lacks data on whether patients had private insurance. However, because the average age of patients in our study was 70 years, and adults over 65 years in Ontario have medication coverage through the provincial government, cost alone is unlikely to explain the low uptake we observed. Our classification of obesity was limited due to the low sensitivity (9%) of ICD-10 codes and the lack of data on BMI in GEMINI. To address these limitations, in addition to ICD-10 codes, we conducted a point prevalence study. This combined approach yielded prevalence estimates for obesity that were consistent with provincial averages for Ontario. We lacked data on when GLP-1RA were added to the hospital formulary at each hospital, affecting our interpretation of the factors that drove low use of GLP-1RA. However, GLP-1RA use in 2021 was low overall, which suggests against hospital-level formulary coverage being the primary reason for low use, since most hospitals in Ontario had added GLP-1RA to their formulary by 2021. We did not exclude individuals with contraindications for GLP-1RA, which include a history of medullary thyroid carcinoma (prevalence: 1 in 150,000 in Canada) or multiple endocrine neoplasia syndrome type 2 (prevalence: 1 in 30,000 - 50,000 in North America).^40^ Given their low prevalence, these contraindications are unlikely to significantly contribute to the low use of GLP-1RA. We did not report on GLP-1RA side effects, which include nausea and vomiting, and it is possible patients were not prescribed a GLP-1RA in hospital because they were acutely unwell. However, that does not preclude its use before discharge when the patient has recovered.

## Conclusion

Clinical guidelines recommend GLP-1RA as a second-line agent for people with T2DM and a first-line agent for people with obesity, but their use among patients who stand to benefit is low. Among over 1.2 million hospitalized adults in Ontario between 2015 and 2022, the prevalence of T2DM and obesity were high, and over 140,000 patients had both T2DM and cardiovascular disease. Hospitalized adults are at high risk for subsequent cardiovascular events, making the inpatient setting an important opportunity to optimize treatment for T2DM and obesity. Our study demonstrates the gap between clinical evidence and clinical practice, and lays the foundation for knowledge translation interventions—including those implemented for hospitalized patients—to improve the uptake of GLP-1RA among those who stand to benefit most.

## Data Availability

All data produced in the present study are available upon reasonable request to the authors.

